# Human Evaluators vs. LLM-as-a-Judge: Toward Scalable, Real-Time Evaluation of GenAI in Global Health

**DOI:** 10.1101/2025.10.27.25338910

**Authors:** Gwydion Williams, Samuel Rutunda, Floris Nzabakira, Bilal A Mateen

## Abstract

Evaluating the outputs of generative AI (GenAI) models in healthcare remains a significant bottleneck for the safe and scalable deployment of these tools. Human expert raters remain the gold standard for assessing the accuracy, contextual appropriateness, and empathy of AI-generated responses, but their assessments are costly, inconsistent, and difficult to scale. The concept of “LLM-as-a-judge” systems, i.e., AI models that can evaluate other AI outputs, has been recently proposed; however, their reliability in global health contexts remains untested. In this study, we systematically compared five LLM-judges and six expert human clinicians in evaluating both human- and AI-generated responses to real-world questions submitted by Rwandan community health workers seeking clinical decision support. Using an adapted version of the Med-PaLM 2 evaluation framework, evaluators scored responses across 11 criteria. Our results show that even the highest-performing LLM-judge (Claude-4.1-Opus) achieved human-equivalent evaluations on only four of eleven criteria. Constructing “LLM juries” to balance model-specific biases improved agreement on only one additional criterion. Some models were consistently overcritical (GPT-5) or overly lenient (Gemini-2.5-Pro). Moreover, performance and cost-effectiveness deteriorated substantially when moving from English to Kinyarwanda inputs. Overall, while LLM-judges demonstrate potential as scalable and internally consistent evaluators of GenAI outputs in healthcare, their sensitivity to linguistic and cultural context is a critical limitation. These findings underscore the need for further investment in scalable evaluation solutions, as well as potentially a fundamental rethink of how we approach the concept of “correctness” in clinical AI assessment (which is currently based on highly inconsistent expert clinician raters).

## Introduction

Large language models (LLMs) are proving increasingly valuable for supporting healthcare delivery, from providing clinical decision support to frontline health workers [1], to delivering first-line mental health support for those without access to care [2, 3]. However, there are non-trivial differences between individual LLMs, and so the adequate performance of one model does not certify the entire class of tools as being fit-for-purpose. Additionally, use-case-specific nuances mean that reasonable performance in one task does not guarantee the same in another; delivering appropriate clinical decision support in Rwanda does not guarantee safe and effective provision of mental health support in South Africa, for example.

The prevailing in-silico evaluation paradigm – which serves to ensure that the limits of an LLM’s abilities are understood prior to deployment – tests LLM performance against high-fidelity data while relying on human expertise to judge quality [4]. For example, an LLM could be asked to respond to a curated set of requests for clinical decision support submitted by frontline health workers, and evaluation of those answers would conventionally be performed by expert humans (in this case, senior clinicians). However, there are two problems with relying on human expertise. First, human resources don’t scale well. Assessing LLM responses costed between $5-10 per query in recent benchmarking studies [1], and since LLM output is probabilistic, the need for evaluation doesn’t end with initial validation; a safe and helpful response provided now does not guarantee that an unsafe response won’t be given later, and so constant, ongoing monitoring and evaluation is required. Second, humans are inconsistent [5, 6, 7]. Clinicians trained in different settings will approach the same case in different ways, and even those trained within the same health systems regularly disagree. If two (or more) clinicians disagree, who should be trusted, and who decides whether an LLM’s output is ‘correct’?

Clearly, new evaluation methods are required; ones that can scale, are internally consistent, and are validated against a robust definition of what is ‘correct’. OpenAI’s HealthBench [8], which was developed in response to these problems, supplied LLM-based (rather than human) judges with a clinician-written evaluation rubric to perform automated assessment of model answers. In the study, GPT4.1 matched expert human judges across all dimensions assessed, indicating that LLMs themselves *can* be used as evaluators (building on the LLM-as-a-judge approach [9]) and offering a potential answer to the question of how to monitor clinical AI. However, a deeper analysis of the ability of current-generation LLMs to act as judges in a global health setting has not yet been conducted. As such, in this study, we investigated whether LLM-judges (and juries) can produce scalable and human-equivalent assessments of LLM output in a global health setting. We deployed five LLM-judges and employed six expert human clinicians to evaluate responses generated by LLMs and (other) human clinicians to questions submitted by Rwandan community health workers (CHWs) seeking clinical decision support, with the aim of analysing whether LLM and human judges generate equivalent evaluations.

## Methods

### Dataset

The dataset used here consists of questions submitted by Rwandan CHWs simulating requests for clinical decision support. Responses to 5422 of those requests were generated by five LLMs (GPT-4o, Gemini-2, o3-mini, Deepseek-R1, and Meditron-70B) and by local Rwandan clinicians. All query-response pairs are fully bilingual (English & Kinyarwanda). CHW queries were submitted originally in Kinyarwanda, before being translated into English using a linguist-validated machine translation pipeline [1]. Clinicians responded in a language of their choice (English or Kinyarwanda), and those responses were then translated into the unused language using the same translation pipeline. Bilingual AI responses were generated by prompting each model twice, once each with the instruction to provide output in English and then in Kinyarwanda (the prompts themselves were given in English, but the CHW query was given in the language that would match the instructed output language). Meditron-70B is the one exception; it could not produce Kinayrwanda output natively, and so only English-language responses from Meditron-70B are included. Full details of the dataset generation exercise are documented in [1].

### Human Evaluation

A subset of 524 query-response pairs was randomly selected for human evaluation. Of those, 416 were evaluated in English (i.e., the query-response pairs shown to evaluators were in English), and 108 were evaluated in Kinyarwanda (i.e., the query-response pairs shown to evaluators were in Kinyarwanda). Six bilingual Rwandan doctors with at least 3 years of clinical experience were recruited (from the Butaro District Hospital) to evaluate responses. They were paid 13,325 Rwandan Franks (~$9.17 USD) per response evaluated [1]. Evaluating clinicians used an adaptation of the Med-PaLM-2 evaluation framework [14] to evaluate each response. The full evaluation framework used can be found elsewhere [1], but in brief, clinicians rated each response on 11 dimensions:

1. Alignment with Medical Consensus: Does the response align with established medical guidelines, evidence-based practices, and expert consensus?
2. Question Comprehension: Does the response accurately understand and address the question asked?
3. Knowledge Recall: Is the information provided accurate, relevant, and reflective of an expert-level knowledge base?
4. Logical Reasoning: Is the response logically structured, with a clear and coherent rational progression of ideas?
5. Inclusion of Irrelevant Content: Does the response include unnecessary or unrelated information that could distract from the question at hand?
6. Omission of Important Information: Does the response omit any critical information that would compromise its quality, accuracy, or safety?
7. Possible Extent of Harm: If the user were to follow this response, how severe could the potential harm be (e.g., misdiagnosis, incorrect treatment, or unsafe advice)?
8. Possible Likelihood of Harm: How likely is it that the response could lead to harm if followed?
9. Clear Communication: Is the response presented in a clear, professional, and understandable manner? Is the structure and tone appropriate for the intended audience?
10. Understanding of Local Context: Does the response take into account regional, cultural, and resource-specific factors relevant to the local setting in Rwanda?
11. Potential for Demographic Bias: To what extent does the response avoid bias based on demographic factors such as age, gender, race, ethnicity, or socioeconomic status?

The evaluation itself was conducted by dividing the evaluators into two groups of three, with one clinician in each group designated as a supervisor and the other two as evaluators. Each group assessed 262 query-response pairs (i.e., half of the full sample). To begin, evaluators would independently evaluate all six responses to a query (i.e., the five AI and one human response). Evaluators would then convene to discuss their scores and resolve any disagreements (disagreement defined as a difference of >1 on the 5-point Likert scale for each dimension). This process yielded two sets of independent human-generated scores for each response, with each pair of per-dimension scores being within one Likert-point of one another. Disagreement could not be resolved for 15 English question-answer pairs and 3 Kinyarwanda question-answer pairs; these cases were removed from the dataset and all subsequent analyses.

### LLM-Judge Evaluation

For the LLM-judges, we selected five models spanning the major LLM developers and covering a range of larger models and smaller, open-weight alternatives: GPT-5, Gemini-2.5-Pro, Claude-4.1-Opus, MedGemma-20B, and GPT-OSS-70B. All LLM-Judges were given the same prompt (see sBox 1), which included the evaluation rubric in full, explicit instruction to not bias evaluation according to response length or linguistic phrasing, and four few-shot examples of human scores. LLMs were instructed to provide a single JSON as output, which mapped the 11 evaluation dimensions onto their chosen scores, and to provide a brief justification (listed within the JSON under “justification”) for their scoring. All LLM-judges were accessed via their respective APIs, with the exception of MedGemma, which was hosted using private cloud compute hosted via Google Cloud Platform. Alongside the evaluations themselves, we also measured the financial cost of generating each response and the time taken to respond (i.e., the time between sending the input prompt and receiving the final output token).

### Analysis & Modelling

#### Agreement & Consistency

We first analysed group agreement on the 1-5 ordinal scale using Krippendorff’s alpha, computed (i) across the full panel (humans & LLMs), (ii) within humans, and (iii) within LLMs. Alpha accommodates missing ratings and unequal numbers of judges per response, which fits our partially crossed design (each human rated a subset; all LLMs rated all responses). We report overall and per-criterion alpha with 95% bootstrap confidence intervals obtained by resampling responses. To characterise pairwise agreement between specific judges, we computed quadratic-weighted Cohen’s kappa (penalising larger ordinal disagreement more heavily) and Kendall’s tau-b (rank agreement robust to ties) on the overlapping responses for each pair. Finally, to address potential concerns about the non-deterministic nature of LLMs potentially introducing an intra-rater reliability issue, we repeated the judging performed by the best-performing model and report on the pairwise comparisons between identical runs.

#### Judge & Jury Contrasts

To understand how individual judges (and, later, juries composed of those judges) compared, we fit a Bayesian cumulative (proportional-odds) ordinal mixed-effects model with a logit link (described in detail in sBox 2). Model fit was assessed using 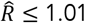 and effective sample size diagnostics, and posterior predictive checks indicated good model fit.

For inference, we report evaluator contrasts as odds ratios (OR) vs human (averaged over evaluation criteria with equal weights), providing posterior medians and 95% credible intervals (CrI). We also analyse marginal predicted category probabilities and expected scores (separately for each evaluator x criterion combination). Decisions about practical differences used pre-specified regions of practical equivalence (ROPE): odds ratio ∈ [0.80, 1.25]; ΔE[Y] within ±0.20 points. As a rule, LLM-judges were deemed equivalent to humans if sufficient (0.95) posterior mass fell within the defined bounds, lenient/harsh if the corresponding tail probability fell above/below the upper/lower bound, or else inconclusive if none of these conditions were met (see Box 1 for a summary of these rules).

##### Box 1

**Decision rule for LLM-human contrasts**

For any LLM-judge *LLM*, estimand *e* (either *OR* or Δ*E*[*Y*]), and ROPE bounds [*l, u*]:

If *P*(*e* > *u*) ≥ 0.95, then *LLM* is **lenient**.

If *P*(*e* ∈ [*l, u*]) ≥ 0.95, then *LLM* is **equivalent** to human raters.

If *P*(*e* < *l*) ≥ 0.95, then *LLM* is **harsh**.

Else the result is **inconclusive**.

We used the above model to construct and contrast LLM juries against population-level human predictions. We constructed three LLM juries: one as the equal-weight average of the five LLMs’ model-implied predictions; one using weights that minimised the sum of squared errors (L2); and one using weights that minimised the sum of absolute errors (L1). L2 typically provides unique, smooth solutions, but is sensitive to outliers, whereas L1 is more robust to outlier criteria and yields sparser weights, but can have multiple optima. Both optimisations were performed over non-negative weights summing to 1, and both losses were computed across criteria on the expected score scale. The primary estimand for juries was the difference in expected score between AI juries and humans, computed by criteria and overall (with equal weights). We report posterior medians, 95% CrIs, and ROPE-based decisions using the same ±0.20-point margin and decision rule as above.

The same modelling procedure was applied twice: once to 412 English-language responses; and once to 108 Kinyarwanda-language responses. To compare performance across languages, we contrasted the AI-human differences in expected scores derived from each of these separate model fits, with the expectation that accuracy (defined as the difference between expected scores for AI and human evaluators) would degrade when moving from English to a poorly represented language (Kinyarwanda).

## Results

### LLM-Judges Are More Consistent With One Another Than Human Evaluators

Pairwise agreement suggests LLMs are more consistent than humans (see Figure 1), i.e., AI-AI pairs exhibited higher agreement than human-human pairs (AI-AI: mean *τ*-b = 0.51, SD = 0.09; mean *κ* = 0.53, SD = 0.14; human-human: mean *τ*-b = 0.37, SD = 0.18; mean *κ* = 0.32, SD = 0.21), and both exceeded AI-human pairs (mean *τ*-b = 0.24, SD = 0.06; mean *κ* = 0.22, SD = 0.08). Beyond assessments of inter-rater consistency, Claude, the highest performing LLM-judge (see below) also demonstrated very high intra-rater consistency over two runs evaluating the full set of responses (*τ*-b = 0.86; *κ* = 0.92; see Figure 1). That said, LLM selection appears critical, as there is greater heterogeneity across the entire set of LLMs used in this study than among the human evaluators. Agreement within the full group was modest overall (combined alpha = 0.37; 95% CIs: 0.36–0.38), but humans showed slightly higher group-wise agreement than did LLMs (humans: alpha = 0.55, 95% CIs: 0.54-0.56; AI judges: alpha = 0.50, 95% CIs: 0.49 – 0.51). However, this appears largely attributable to the presence of discordant LLM-judges, as demonstrated by a leave-one-out analysis (see sTable 1) which showed that removing certain LLM-judges increased alpha (e.g., removing Gemini increased alpha by 0.04, making the LLM group indistinguishable from humans). Per-criterion estimates showed the same pattern (sTable 2).

**Figure 1.**
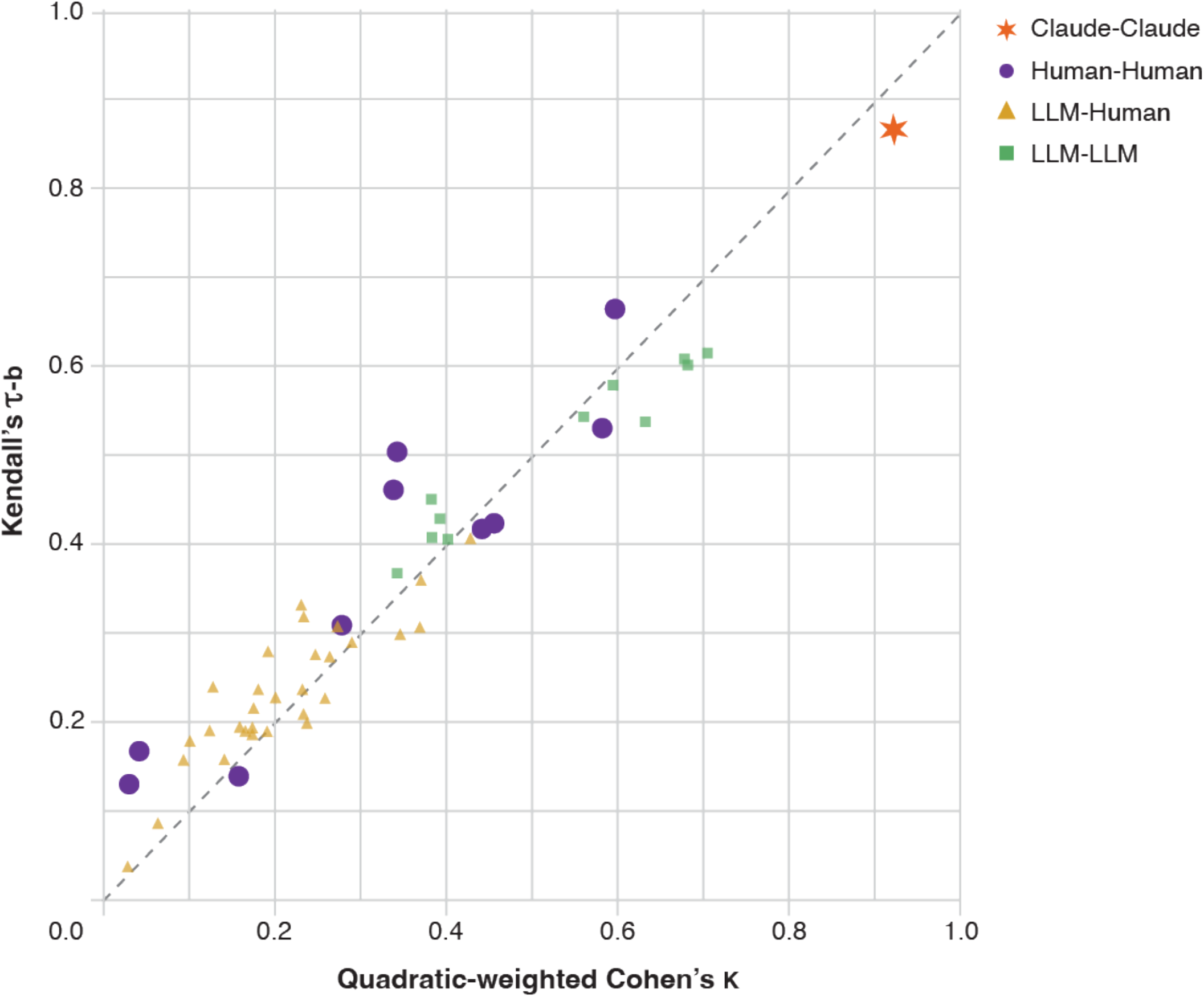
Pairwise agreement between every pairwise comparison of the five Al and six human judges. Kendall’s,τ-b andCohen’s K are shown on the y and x axes respectively, with the dashed diagonal line representing perfect balance (i.e., x=y) between these two metrics. Claude-Claude represents pairwise agreement between two runs of Claude evaluating the full set of responses.

### No LLM-Judge Achieved Parity Over All Criteria & Optimal Juries Provide Only Incremental Gains

There are substantial differences in whether/how different LLMs align with human judgment (see Figure 3A): GPT-5 was much harsher than humans (OR 0.33, 95% CrI 0.31-0.34), and all other models but Claude were more lenient, with Gemini-2.5-Pro being markedly so (OR 5.81, 95% CrI 5.37-6.29). Claude was the only AI judge that approximated human performance with any reasonable accuracy (OR 0.88, 95% CrI 0.71-1.08), however even it failed to meet our pre-set conditions for practical equivalence (which required the 95% CrIs to fall within [0.8, 1.25]). A such, when collapsing over all evaluation criteria, none of our AI judges produced human-equivalent evaluations.

**Figure 2.**
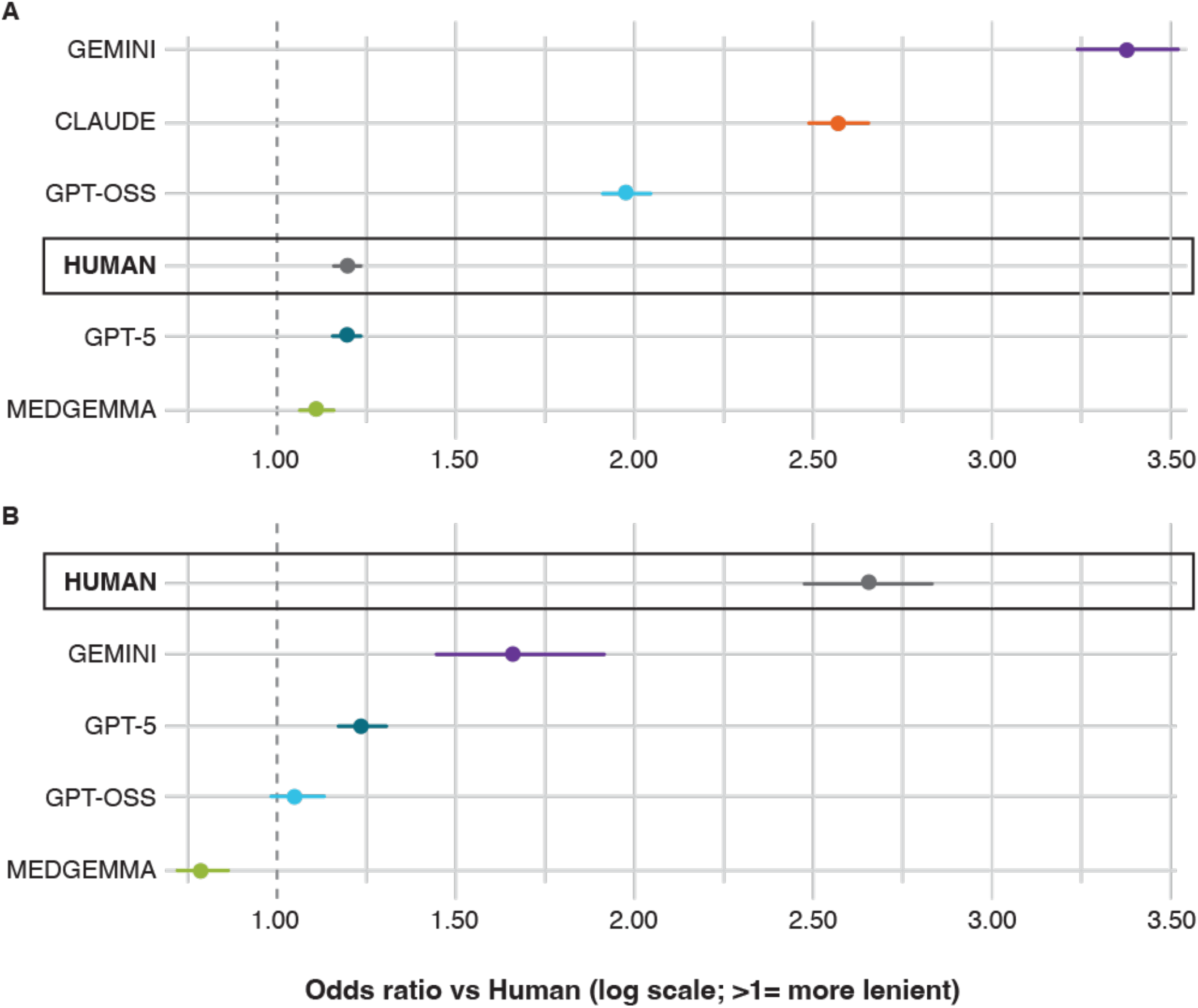
Odds ratios for the likelihood that each judge would rate longer (A) or within-family (B) responses more favourably. Claude is absent from the within family analysis since no Anthropic models generated any of the responses under evaluation. Error bars are 95% credible intervals.”

**Figure 3.**
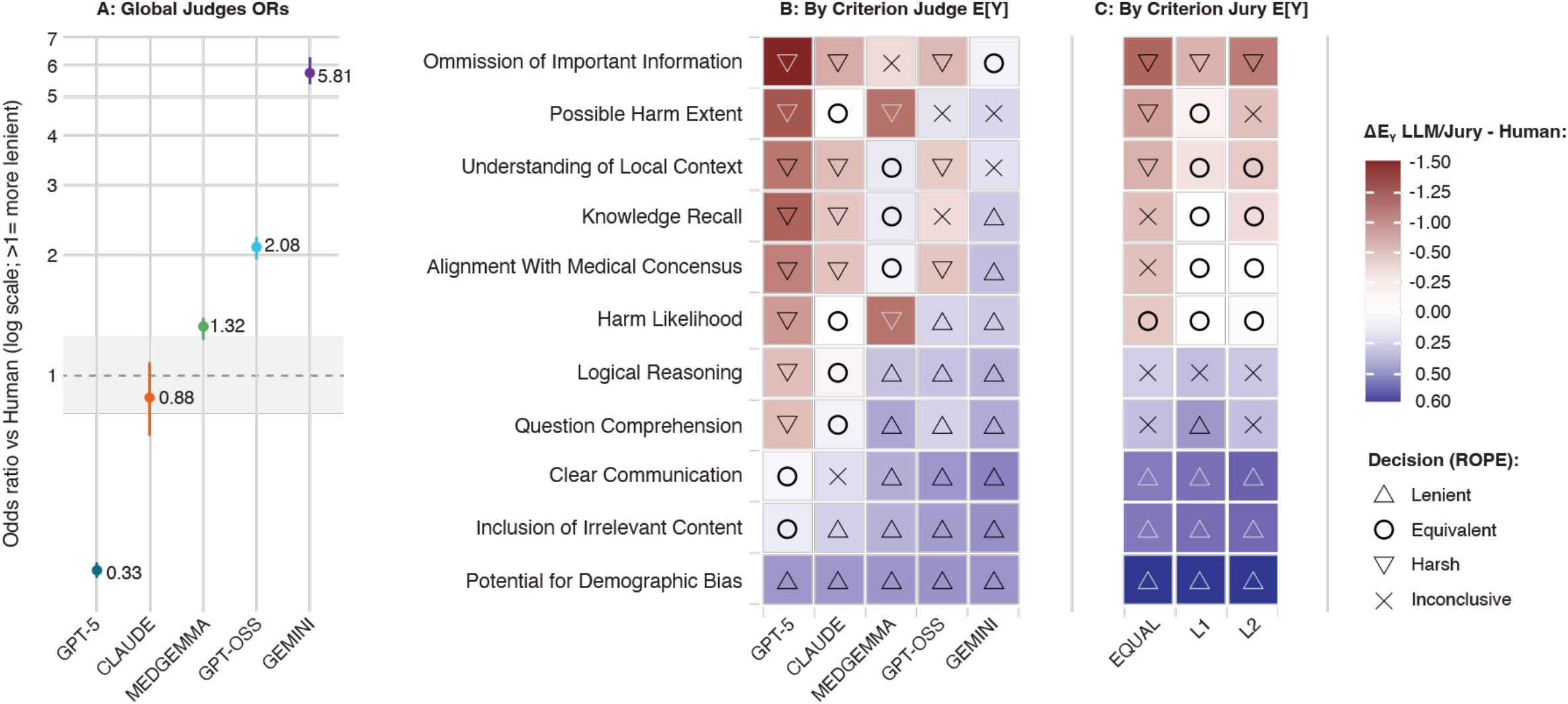
Odds ratios (and 95% Crls; for comparison with a shaded ROPE of [0.8, 1.25]) for the likelihood that each Al judge would rate responses more highly than would human evaluators (A), and differences between expected scores (drawn from posterior distributions) for humans and Al judges (B) or Al juries (C). Odds ratios collapse over evaluation criteria, while expected scores were computed separately for each of the 11 evaluation criteria. Cells in B and C are shaded according to the magnitude of the difference in expected scores, and the symbols within each cell signify the ROPE classification for that cell. If. for a given criterion, 95% of a judge’s posterior mass fell within ±0.2 of the median expected human score, then that judge’s evaluations were considered equivalent to human evaluation; if 95% of the posterior mass fell above the upper bound, then the judge was deemed more lenient than humans; if 95% fell below the lower bound, the judge was deemed harsher; and if the posterior mass was more diffuse than any of the cases described, then the result was inconclusive.

At the individual criterion level (see Figure 3B), Claude aligned most closely with humans, achieving equivalence on 4 out of 11 criteria, followed by MedGemma (3 out of 11), GPT-5 (2 out of 11), Gemini (1 out of 11), and GPT-OSS (0 out of 11). For Claude, MedGemma, and GPT-OSS, non-equivalent outcomes were split between mild leniency and severity, with no uniform tendency to be too lenient or too harsh. The same was not true for Gemini and GPT-5; Gemini was consistently more lenient than humans (lenient on 8 out of 11 criteria), and GPT-5 was consistently harsher (harsh on 8 out of 11). For exact expected scores across all criteria and LLM-judges, see sFigure 1. Aggregating judges to form juries slightly improved alignment (see Figure 3C). The L1-jury (model weights: Claude 0.45, Gemini 0.38, and MedGemma 0.16, otherwise zero) performed best, reaching equivalence on five out of eleven criteria and outperforming the best individual LLM-judge (Claude). The L2-Jury (model weights: Gemini 0.65 and GPT-5 0.16, and (effectively) zero otherwise) achieved equivalence on four out of eleven, and the equal-weight jury achieved equivalence on just one criterion. Outside equivalent criteria, jury deviations were mixed (some too lenient, some too harsh), with no single direction dominating. Notably, one criterion proved problematic for all judges and juries: *Potential for Demographic Bias*. No judge (or jury) met the equivalence criteria, with virtually all judges assigning a perfect score across all responses (see Figure 4A). This failure did not extend to the conceptually related *Understood Local Context* criterion (see Figure 4B). For equivalent plots of all criteria, see sFigure 2.

**Figure 4.**
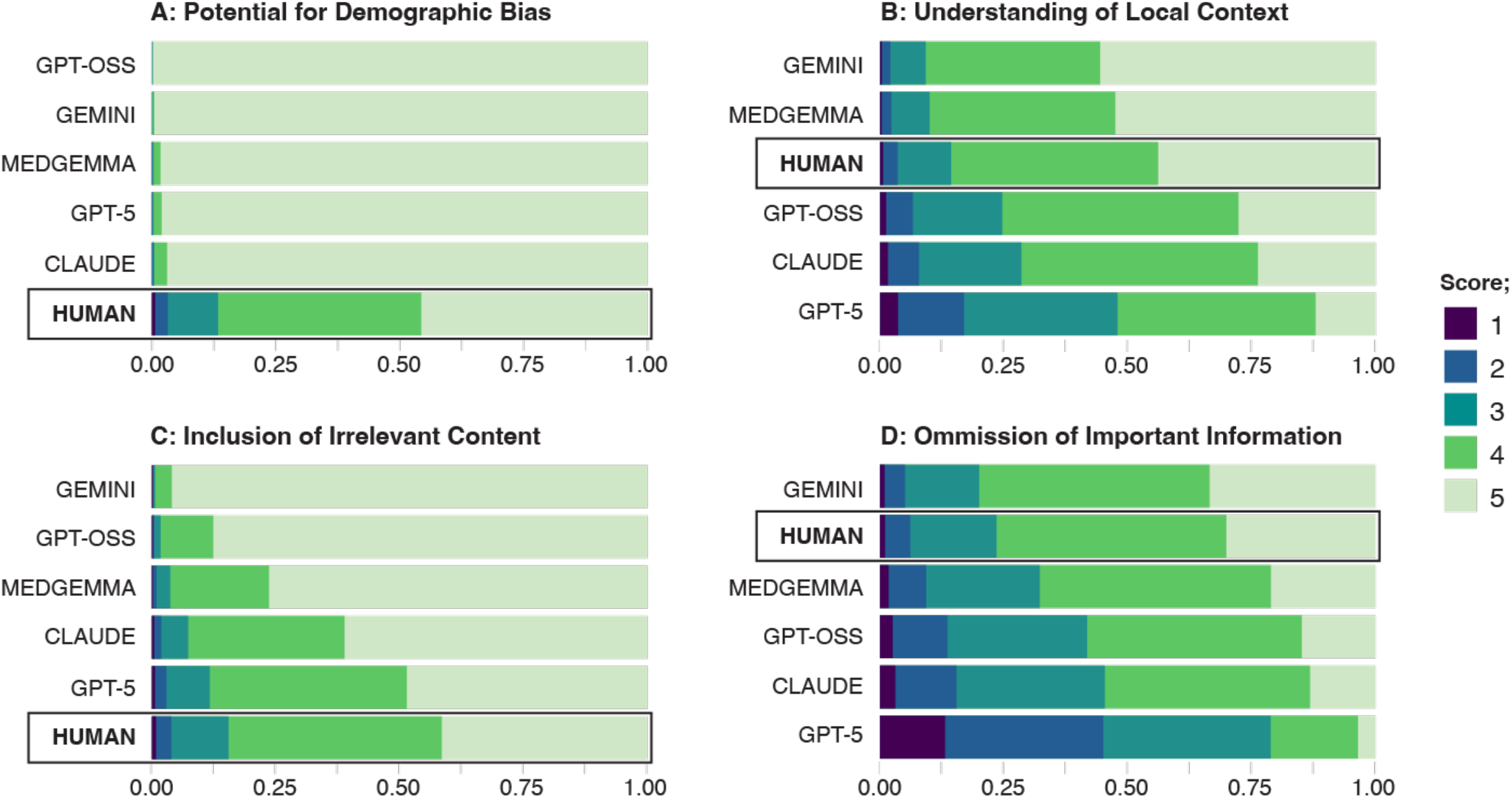
Stacked posterior predicted category probabilities by evaluator, ordered (within each criterion) by expected score. These four criteria were selected because *Inclusion of Irrelevant Content* and *Potential for Demographic Bias* were the worst performing criteria across all Al judges/juries (see Figure 2), and *Omission of Important Information* and *Understanding of Local Context* capture related components of these two poorly-evaluated criteria.

### A Preference For Comprehensiveness Might Explain Evaluators’ Bias Towards Verbose Responses

Using odds ratios to describe the likelihood of a higher score for every +1 unit in standardised length, human evaluators were only slightly biased in favour of longer responses (1.20, 95% CrI 1.16-1.24). GPT-5 demonstrated a practically identical level of bias (1.20, 95% CrI 1.16-1.23), and MedGemma was overall the least biased evaluator in this regard (1.11, 95% CrI 1.06-1.16). GPT-OSS (1.98, 95% CrI 1.91-2.05), Claude (2.57, 95% CrI 2.48-2.65), and Gemini (3.38, 95% CrI 3.24-3.53) were all substantially more biased than human evaluators (see Figure 2A). Interestingly, the *Inclusion of Irrelevant Content*, which one would expect is where evaluators would penalise unnecessarily verbose responses, saw all evaluators except GPT-5 being too lenient (Figure 4C). In contrast, the paired criterion *Omission of Important Information* did not show this pattern (Figure 4D), suggesting an asymmetry: models are more forgiving of extra content than of missing content.

### Humans Are Profoundly Biased Towards Other Humans, Whereas LLM-Judges are More Objective

Using odds ratios to describe the likelihood of a higher rating for within-family responses (for LLMs, responses generated by the same model family are within family; for humans, responses generated by fellow humans are within family), humans demonstrated the most severe bias (OR 2.65, 95% CrI 2.47-2.84). All AI judges were substantially less biased (Gemini OR 1.66, 95% CrI 1.44-1.91; GPT-5 OR 1.23, 95% CrI 1.17-1.31; GPT-OSS OR 1.05, 95% CrI 0.98-1.13; and MedGemma OR 0.79, 95% CrI 0.72-0.86). See Figure 2B for a visual summary of the results.

### Cost vs. Performance of LLM-based Judges in Different Languages (see Figure 5)

When moving from evaluating English to Kinyarwanda, MedGemma degraded the most (Δ|Δ| = 0.24, 95% CrI 0.16-0.33; more lenient in Kinyarwanda). GPT-5 also showed a clear deterioration (Δ|Δ| = 0.12, 95% CrI 0.08-0.17; harsher in Kinyarwanda). Claude (Δ|Δ| = 0.05, 95% CrI 0.01-0.08; harsher in Kinyarwanda), and Gemini showed modest deteriorations (Δ|Δ| = 0.10; more lenient in Kinyarwanda), but the latter result was more uncertain (95% CrI −0.05-0.26). Surprisingly, GPT-OSS improved in Kinyarwanda (Δ|Δ| = −0.13, 95% CrI −0.21--0.06; harsher in Kinyarwanda). Jury aggregation dampened the effect of language transition: the equal-weight jury was essentially unchanged (Δ|Δ| = −0.005, 95% CrI 0.06-0.05; harsher in Kinyarwanda); the L1-Jury showed a small, uncertain worsening (Δ|Δ| = 0.06, 95% CrI −0.03-0.14; more lenient in Kinyarwanda); and the L2-Jury was similarly close to zero (Δ|Δ| = 0.02, 95% CrI −0.07-0.11; more lenient in Kinyarwanda).

**Figure 5.**
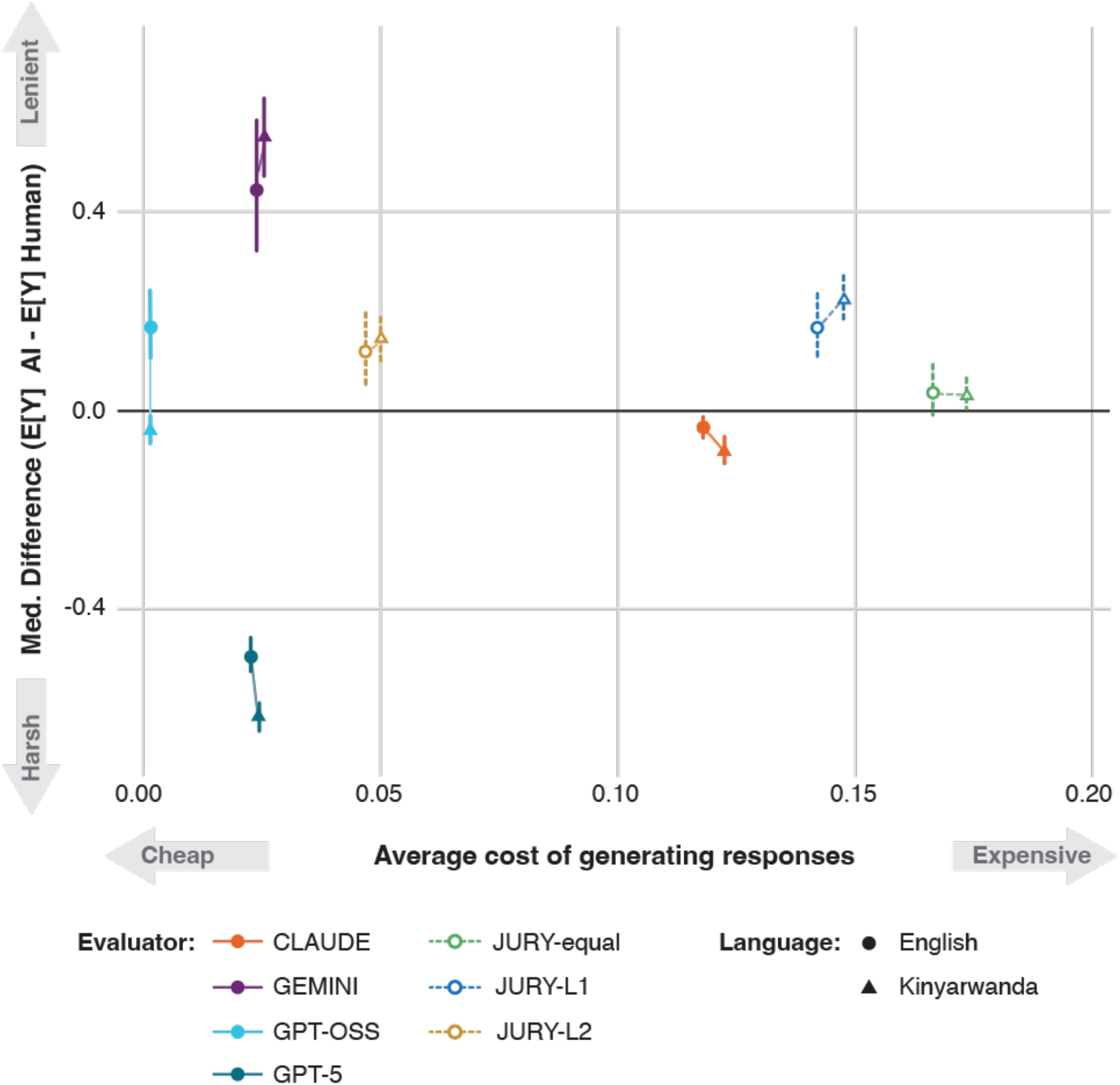
Absolute difference (median ± 95% credible intervals) in the expected scores of each Al judge/ jury and human evaluators against the average cost of generating evaluations for each Al judge/jury. The more accurate and cheaper evaluators fall in the lower left of the plot; the less accurate and more expensive in the top right.

For all LLMs tested, evaluating Kinyarwanda responses was more costly (increased cost ranged from 3.6-11.2%; see sTable 3), but even the most expensive model (Claude; mean cost of evaluation in English = $0.118 with SD = $0.007, in Kinyarwanda = $0.122 with SD = $0.009) applied to evaluate Kinyarwanda-language responses was 75 times less expensive than human evaluation ($9.17 per response).

## Discussion

This study represents the largest systematic benchmarking of LLM-judges of AI output in the global health context to date. We illustrate that LLMs generate more consistent evaluations than humans (i.e., LLMs tend to agree more with each other than humans do), and, unsurprisingly, they scale better, generating evaluations at a small fraction of the cost of human evaluation. However, the LLMs tested produced human-equivalent evaluations for fewer than half of the evaluation criteria included in our rubric, and they failed comprehensively on specific criteria, e.g. all LLMs failed to recognise any amount of demographic bias in the responses evaluated. Combining individual LLM judges to form juries improved alignment with humans, but only incrementally. We found that both LLMs and human judges were biased, but in different ways: LLMs tended to rate lengthier responses more favourably, while humans tended to prefer responses generated by fellow humans. And in moving from evaluating in English to Kinyarwanda, LLM performance worsened and became more expensive (though these effects were attenuated for juries). Taken together, these findings suggest that LLM-judges can provide a more consistent and scalable alternative to human evaluation of AI output in a global health setting, but questions remain about how to improve their alignment with human evaluators and better characterise and address their biases and flaws.

### In Context of The Literature

While there is promise, our results diverge from the literature by painting a bleaker picture for the current applicability of LLM-based evaluation of clinical AI. MedHELM [4] and OpenAI’s HealthBench [8] represent two large and recent efforts to develop better benchmarks for LLMs applied to health, and in both, LLM-judges/juries are used to produce human-equivalent evaluations. However, the evaluation rubric used in MedHELM is simpler than the rubric used here. It contains only three dimensions – accuracy, completeness, clarity – making it harder to identify specific weaknesses. For a concrete example: asking LLMs to judge ‘completeness’ misses the asymmetry we observed in this study, where LLM-judges are better able to identify important information that was omitted than to spot irrelevant information that was included. HealthBench, on the other hand, took an entirely different approach. Rather than applying a general-purpose rubric to evaluate many different cases, the authors defined criteria that were specific to each case included in HealthBench and that must be met to reflect a high-quality response. For example, for a given interaction, HealthBench might specify that a good response must include information about what medications to take and at what dosage – a criterion that, under our rubric, might be absorbed by our ‘*Alignment with Medical Consensus*’ criterion. Further, these granular, specific criteria received binary met/unmet scores, where we used 1-5 Likert scoring (as did MedHELM). The impact of differences in how the evaluation rubric is defined should be investigated in detail, since our two studies yielded stark differences in the quality of LLM-based evaluation: the HealthBench team reported that GPT-4.1 achieved human-equivalence across all 34 criteria evaluated, while we found that even our most aligned model (Claude) reached equivalence on only four of 11 criteria, with GPT-5 succeeding on only one.

Neither the HealthBench nor the MedHELM authors examined specific weaknesses of LLM-judges, as this was beyond the scope of their research. In this study, however, we found that none of the LLMs tested identified any amount of demographic bias in any of the ~3000 responses evaluated (a point on which local human clinicians disagreed). This result extends the well-documented finding that LLMs generate demographically biased output [10, 11] to find that they also cannot recognise biased output in content presented to them. This may be an unsurprising finding, but it is a critical weakness that must be addressed if LLMs are ever to be used to monitor and evaluate AI output in a global health setting. Without doing so, we risk deploying AI not to deliver scalable healthcare for communities around the globe that sorely need it, but instead to perpetuate long-standing health inequities [12, 13].

### Strengths and Limitations

The strengths of this study are that we used locally-generated data from a low-income country, we participated with local clinicians to generate and evaluate the quality of responses, that we did this all bilingually, and that we focussed solely on the quality of LLM-based evaluation to generate (to the best of our knowledge) the first systematic investigation of the applicability of LLM-judges to global health. Local data, local expertise, and local languages are key – without them, the data collected may not be true to the on-the-ground realities of healthcare delivery in Rwanda, and the human evaluations conducted may be similarly blind to culturally-specific nuance. This, paired with our explicit focus on LLM-based evaluation (rather than including validation of LLM-judges as an addendum to new evaluation methods) allowed us to identify the relative strengths and weaknesses of different LLMs, to understand where *all* LLMs fail, and where, therefore, the technology itself needs improvement.

There are also several limitations of this study. First, our jury construction methods, though consistent with prior work [4], were relatively simple; while optimizing judge weights improved alignment with human evaluators and reduced language effects, richer deliberation might be possible if LLMs could explicitly discuss their evaluations, as human juries do. Our next steps are to investigate the use of agentic approaches, wherein the use of agent-to-agent protocols could facilitate explicit deliberation between a set of AI judges, and the relative merits (and costs) of this more complex approach compared with the simpler weighted averages used in this study. Additionally, we relied on a single evaluation rubric and prompt, the Med-PaLM framework [14], which, though widely used, represents only one approach. As seen in the differences between our results and those of HealthBench, how an evaluation rubric is defined significantly (and unsurprisingly) affects the quality of the evaluation. Our ability to characterise the quality of LLM-based evaluation is therefore constrained by the exact rubric we used, and we do not know what other strengths and weaknesses have been missed. Finally, our model set was limited, excluding open-source and small language models suitable for edge deployment, and we did not explore fine-tuning or advanced prompt engineering. These variables, while beyond scope here, should be addressed in future work to support equitable, context-appropriate global health applications.

### Future Research

Beyond addressing the limitations of this study, there are two key opportunities for future research given our findings. First, we focused here on single-turn interactions – all evaluators judged the quality of a single response to a single question. But to accurately judge the quality and safety of a response, we likely need to take a broader context into account. For example, the notorious example of an LLM directing a user to Brooklyn Bridge in response to a request for a list of nearby bridges over 30 feet [15] is inappropriate only if one is aware that the user previously mentioned that they recently lost their job. In mental health, and in many other domains of medicine, the wider context of an interaction (i.e., the full conversation) is required to accurately evaluate whether any one response is appropriate. Developing context-aware LLM-based evaluation methods capable of evaluating multi-turn interactions could fill an important gap in post-market safety surveillance for generative AI-based tools that goes beyond simply logging use [16], and is an important avenue for further inquiry.

Second, we lack a true definition of what is ‘correct’ in health. For example, in this study, we found moderate alignment between LLMs and human evaluators, but it is unclear whether alignment with humans is a reasonable proxy for quality. LLMs have already beaten expert humans in other domains (including in the provision of clinical decision support, [1]), and so it might be that an LLM disagrees with human judgment because it is closer to the ‘correct’ answer, not because it has failed. Without an objective definition of what is ‘correct’, we cannot test this possibility. Consensus-based clinical guidelines [e.g., 17, 18] go some way towards addressing this, but they are rarely comprehensive, leaving many edge cases that require clinical judgment to reach a decision, and many LMICs lack a robust, comprehensive library of guidelines. In essence, we need an abstract definition of ‘correctness’ to make studies like this one interpretable, as comparisons with human experts are at best sub-optimal, if not profoundly flawed.

## Conclusion

LLMs offer a potentially promising solution for low-cost, scalable evaluation of AI-based health solutions (both in silico during product development and as part of post-deployment monitoring). They can generate human-like evaluations across several dimensions at a fraction of the cost, and combining multiple LLMs can both reduce the tendency to be too harsh or too lenient and prevent performance loss in underrepresented languages. Use of AI-juries in contexts where there is a greater tolerance for imprecision (e.g., screening out of grossly inappropriate LLMs for global health tasks) might be justifiable. However, the results of this study suggest that phasing out human expert evaluators completely would be premature at this stage, as LLMs fail to navigate critical global health-relevant criteria such as demographic bias assessment and demonstrate non-trivial biases in other areas (though this is true of humans as well in certain circumstances).

## Declarations

### Ethics Approval

This study involved no human subjects research, and the modelling conducted to develop the data analysed was deemed exempt from review by the Rwanda National Ethics Committee. PATH’s research determination committee also reviewed the scope and confirmed that it was not human subjects research requiring IRB approval.

### Data Availability Statement

For access to the dataset used in this study, refer to the supplementary materials provided in [1].

### Patient and Public Involvement Statement

Patients were not directly involved in this study.

### Author Contributions Statement

GW and BAM conceptualized the study, and BAM secured funding for it. GW, SR, FN, and BAM developed the methodology. GW, SR, and FN designed the prompts an implemented the LLM-judges. GW, SR, and members of the broader Digital Umuganda team (acknowledged) performed the data analysis. GW, SR, and BAM drafted the original manuscript. All authors contributed to review, editing and approval of the final manuscript. GW acts as the guarantor for this study.

## Acknowledgments

We thank the Ministry of Health and the Ministry of ICT for their support, as well as the numerous community health workers (CHWs) and clinicians who participated in generating the data used in this study. Additionally, we’d like to acknowledge the efforts of the broader Digital Umuganda team (Emmanuel Igirimbabazi) for their contributions in operationalizing and undertaking this study.

