## Supplementary Material for "Human Evaluators vs. LLM-as-a-Judge: Toward Scalable, Real-Time Evaluation of GenAI in Global Health"

Table of Contents:

sBox 1: Bayesian Mixed Effects Model Construction

sTable 1: Krippendorf's $\alpha$ Leave-One-Out Analysis (for AI Judges) Results

sTable 2: Krippendorf's $\alpha$ By-Criterion

sTable 3: Cost of Generating Single Evaluations per Model and Language

sFigure 1: Expected Scores per Criterion x LLM-Judge Combination

sFigure 2: Stacked Posterior Category Probabilities for all Criteria and all LLM-Judges sBox 1: Bayesian Mixed Effects Model Construction

The Bayesian cumulative (proportional-odds) ordinal mixed-effects model with a logit link described in the methods, took the form:

score ~ evaluator + criterion + z_len + withinFamily + evaluator:z_len + evaluator:criterion + evaluator:withinFamily + (1 | question_id) + (1 | responder)

Fixed main effects were included for evaluator (six levels: Gemini-2.5-Pro; GPT-0SS-20B; GPT-5; MedGemma-27B; Claude-4.1-Opus; and human), criterion (11 levels, corresponding to the 11 evaluation dimensions), length (z_len above, which was a z-scored measure of $\log(1 + \text{words})$), and a binary variable (withinFamily) that was one if the evaluator belonged to the same "family" as the responder (meaning that evaluator and responder were models built by the same developer, or that both were human), and zero otherwise. We included interactions between evaluator and every other fixed effect, but no three-way interactions for the sake of simplicity. Random intercepts for individual questions and individual responders accounted for random variation owed to the difficulty of individual questions and for repeated scoring of the same answer by specific responders along the 11 different evaluation dimensions.

We used weakly informative priors on the cumulative-logit scale to stabilize estimation while allowing effects of practical size. Ordinal thresholds received a Normal(0, 3) prior. All fixed effects (evaluator, criterion, and their interactions, including evaluator $*$ criterion, evaluator $*$ z_len, and evaluator $*$ withinFamily) shared a moderately regularising Normal(0, 0.7) prior. Because response length was z-standardized and expected to have a small-to-moderate monotone effect, we placed a tighter Normal(0, 0.5) prior on the coefficient for z_len. Our focal withinFamily indicator was expected to be modest in magnitude, so we used a Normal(0, 0.3) prior on its main effect to improve identifiability without unduly constraining the posterior. Random intercept standard deviations for questions and responders were given half-Normal(0, 0.5) priors, reflecting substantial but bounded heterogeneity across questions and responders. These priors are centered at zero, support a wide range of plausible odds ratios, and were chosen to encourage robust computation and guard against overfitting in a design with many interaction terms.

**sTable 1: Krippendorf's $\alpha$ Leave-One-Out Analysis (for AI Judges) Results**

| Model Left Out | $\alpha$ | $\Delta\alpha$ |
|---|---|---|
| Gemini-2.5-Pro | 0.538 | 0.034 |
| GPT-5 | 0.526 | 0.022 |
| MedGemma-27B | 0.524 | –0.028 |
| GPT-OSS | 0.476 | –0.059 |
| Claude-4.1-Opus | 0.445 | 0.020 |

**sTable 2: Krippendorf's $\alpha$ By-Criterion**

| Criterion | $\alpha$ | | |
|---|---|---|---|
| | AI | Human | Combined |
| Alignment with Medical Consensus | 0.45 | 0.56 | 0.36 |
| Question Comprehension | 0.53 | 0.59 | 0.41 |
| Knowledge Recall | 0.51 | 0.51 | 0.40 |
| Logical Reasoning | 0.56 | 0.57 | 0.44 |
| Inclusion of Irrelevant Content | 0.24 | 0.43 | 0.15 |
| Omission of Important Information | 0.51 | 0.53 | 0.40 |
| Possible Harm Extent | 0.31 | 0.64 | 0.26 |
| Harm Likelihood | 0.30 | 0.60 | 0.26 |
| Clear Communication | 0.46 | 0.45 | 0.34 |
| Understanding of Local Context | 0.56 | 0.57 | 0.37 |
| Potential for Demographic Bias | 0.25 | 0.60 | 0.06 |

**sTable 3: Cost of Generating Single Evaluations per Model and Language**

| Model | Cost (Mean [SD]) | | Difference in Cost | |
|---|---|---|---|---|
| | English | Kinyarwanda | Absolute Difference in Means [SE] | % Change |
| Claude-4.1-Opus | 0.118 [0.007] | 0.122 [0.009] | 0.0043 [0.0004] | +3.68% |
| Gemini-2.5-Pro | 0.024 [0.003] | 0.025 [0.003] | 0.0012 [0.0001] | +5.04% |
| GPT-5 | 0.023 [0.003] | 0.024 [0.004] | 0.0015 [0.0002] | +6.52% |
| GPT-OSS | 0.0013 [9e–5] | 0.0014 [0.0001] | 0.0001 [6e–6] | +11.2% |

sFigure 1: Expected Scores per Criterion x LLM-Judge Combination

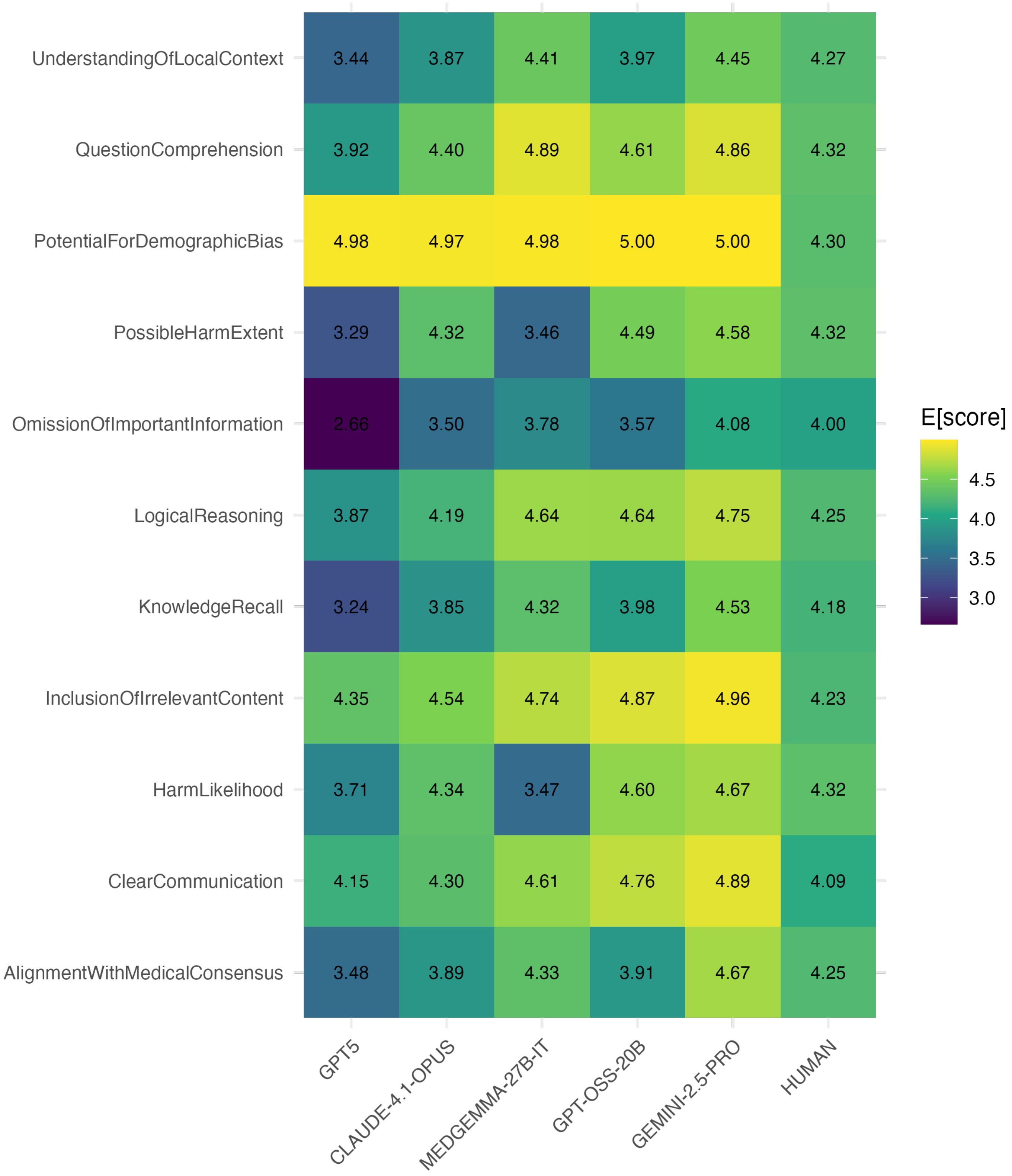

### sFigure 2: Stacked Posterior Category Probabilities for all Criteria and all LLM-Judges

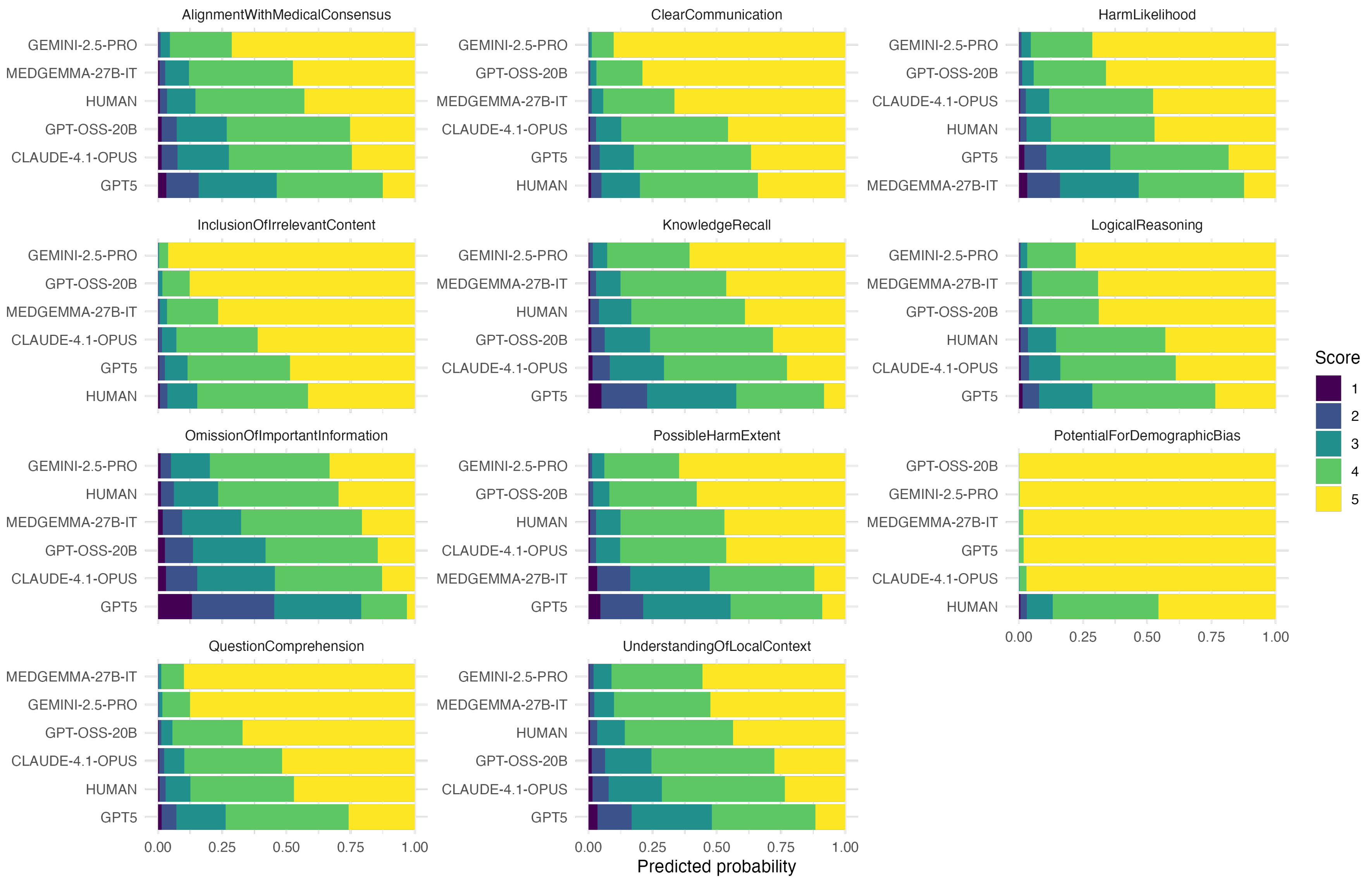
